# Uneven Recovery of Exercise Training in US Adults With Cardiovascular Risk Factors or Disease Following the COVID-19 Pandemic

**DOI:** 10.64898/2026.10.09.26363506

**Authors:** D. August Oddleifson, Jiacheng Zu, Rishi K. Wadhera

**Author notes:** **Corresponding author:** Rishi K. Wadhera, MD, MPP, MPhil, Section of Health Policy, Richard A. and Susan F. Smith Center for Outcomes Research, Beth Israel Deaconess Medical Center, 375 Longwood Ave, 4th Floor, Boston, MA, 02215, USA.

## Abstract

**Introduction:** The COVID-19 pandemic led to significant disruptions in cardiovascular care and reductions in physical activity, but little is known about how exercise patterns changed after this period. This study assessed changes in weekly aerobic and strength training activity between 2020 and 2022 amongst adults aged 18 years or older who reported one or more cardiovascular diseases or risk factors.

**Methods:** This study used National Health Interview Survey data from 2020 and 2022. Adjusted differences for moderate-to-vigorous physical activity (MVPA) were calculated using a linear regression model and for strength sessions using a Poisson regression, adjusting for sex and age. Changes in these outcomes were then stratified by age and sex subgroups.

**Results:** The weighted study population included 182,783,008 (standard error, ±3,334,337) adults in 2020 and 185,388,060 (SE, ±3,285,161) adults in 2022. Between 2020 and 2022, the overall weekly MVPA increased from 244.2 (SE, ±4.5) minutes to 266.3 (SE, ±5.7) minutes (adjusted difference, +22.1 [SE, ±6.92] minutes). There were no statistically significant changes in the number of strength session per week overall or among sex and age subgroups. Across sex and age subgroups, increases in weekly MVPA were entirely concentrated among males and younger adults.

**Conclusion:** The recovery from pandemic era reductions in aerobic and strength exercise training was uneven amongst demographic subgroups, which aligns with concurrent persistent disruptions in cardiovascular care.

## Introduction

The COVID-19 pandemic led to severe disruptions in cardiovascular care, as well as major behavioral and social shifts in the United States.^1,2^ Daily routines were upended, with adults spending more time working from home and less time commuting.^3^ Prior research suggests that the early pandemic was associated with decreased physical activity for people with and without chronic diseases globally, likely due to restrictions from lockdown policies.^4^ However, little is known about how exercise patterns changed after this period, particularly among people with cardiovascular risk factors or disease. Therefore, we assessed national changes in weekly aerobic activity and strength training in this population, both overall and across subgroups (age, sex) between 2020 and 2022.

## Methods

We used National Health Interview Survey (NHIS) data from 2020 and 2022. NHIS is an annual survey of the noninstitutionalized, civilian US population that interviews a representative sample of adults regarding health, health care access, and health behaviors. Respondents report the frequency, duration, and type (light, moderate, and vigorous) of aerobic activity along with the number of strength sessions per week. NHIS weights are adjusted for non-response and under-coverage to generate nationally representative estimates of the US population. Survey design changes in 2020 prohibited the use of pre-pandemic data.

Our study population included adults aged 18 years or older who reported one or more cardiovascular diseases (coronary artery disease, angina pectoris, heart attack, stroke) or risk factors (hypertension, diabetes, hyperlipidemia, obesity, cigarette use). We estimated total weighted moderate-to-vigorous physical activity by first multiplying the reported frequency and duration of both activity types and then adding the totals. Minutes of vigorous physical activity were multiplied by two to account for intensity, as is consistent with AHA physical activity recommendations and prior studies ^5^. We then calculated adjusted differences between 2020 and 2022 for moderate-to-vigorous physical activity (MVPA) using a linear regression model and for strength sessions using a Poisson regression. Our models adjusted for sex and age. We then evaluated changes in these outcomes from 2020 to 2022 stratified by age and sex subgroups.

Significance was considered p<0.05. IRB approval was not obtained from BIDMC because we used publicly available deidentified data, as per institutional policy.

## Results

The weighted study population included 182,783,008 (standard error, ±3,334,337) adults in 2020 and 185,388,060 (SE, ±3,285,161) adults in 2022. Demographic characteristics were similar across study years. In 2020, 50.4% of the study population was female. The age distribution was 36.7% between 18 to 44 years, 36.1% between 45 and 64 years, and 27.2% over 65 years (**Table**).

Between 2020 and 2022, the overall weekly MVPA increased from 244.2 (SE, ±4.5) minutes to 266.3 (SE, ±5.7) minutes (adjusted difference, +22.1 [SE, ±6.92] minutes) (**Figure**). Among young adults ages 18-44, weekly MVPA significantly increased from 289.7 (SE ±8.6) minutes to 316.9 (SE, ±10.7) minutes (adjusted difference, +27.6 [SE, ±14.0] minutes). These patterns were similar among middle-aged adults ages 45-64, who increased from 231.5 (SE ±6.8) minutes to 255.8 (SE ±8.4) minutes (adjusted difference, +23.9 [SE, ±10.6] minutes). However, older adults aged 65+ experience no significant change in MVPA. Across sex subgroups, increases in weekly MVPA were entirely concentrated among males (292.3 [SE, ±7.4] minutes to 328.1 [SE, ±9.0] minutes, adjusted difference +36.5 [SE, ±11.5] minutes).

There were no statistically significant changes in the number of strength session per week overall or among sex and age subgroups **(Figure)**. Males (1.36 [SE, ±0.03] sessions/week) and young adults ages 18-44 (1.44 [SE, ±0.04] sessions/week) had the highest number of strength sessions per week in 2020.

## Discussion

This study found that aerobic exercise increased by an average of 22 minutes per week between 2020 and 2022 among adults with cardiovascular risk factors or disease. In contrast, there were no changes in strength training. Males and young adults had the greatest increases in aerobic physical activity. No demographic subgroup showed a decline in either aerobic or strength physical activity.

The modest increases in physical activity observed in this study may represent an early recovery from the decline seen during the pandemic. This trend may have provided some limited benefit in offsetting the disruptions in cardiovascular care that persisted after 2020.^2^ However, it is concerning that our results suggest that this recovery was uneven, with gains entirely concentrated amongst males and young adults. This likely represents underlying differences in the persistence of effects of the pandemic on exercise patterns. For example, older adults may rely more on caregivers, healthcare providers, or their community for physical activity. These relationships may have been more vulnerable to disruption during the pandemic and slower to restore. Additional research is needed to better understand the sociodemographic drivers of these observed subgroup differences.

Strength training also warrants renewed clinical and investigative focus given the observed stagnation. Recent research suggests that strength training improves many traditional cardiovascular risk factors, including blood pressure, glycemia, lipid profiles, and body composition, and is associated with a significant reduction in all-cause and cardiovascular mortality.^5,6^ Unfortunately, less than a third of US adults meet the recommended 2 days/week of strength training.^6^

Our results support population- and individual-level interventions to promote physical activity, a modifiable and accessible means for reducing cardiovascular risk and preventing cardiovascular disease^5^. Public health campaigns can provide education on the health benefits, deliver guidance on how to exercise, and promote local community-based events and activities. In the clinical environment, cardiologists can assess exercise patterns, offer solutions to perceived barriers to exercise, and refer eligible patients to in-person or virtual cardiac rehab, a chronically underutilized resource.

Limitations of this study included the use of self-reported exercise data which may be subject to recall bias, the inability to assess trends prior to 2020 due to changes in NHIS survey design, and potential differential survey participation across demographic groups during the pandemic.

In conclusion, this study demonstrates uneven recovery from pandemic era reductions in aerobic and strength exercise training, which aligns with concurrent persistent disruptions in cardiovascular care. Targeted public health and clinical interventions are needed to improve adherence to exercise guidelines, a crucial strategy in ongoing efforts to prevent cardiovascular disease.

## Data Availability

All data produced are available online at the National Health Interview Survey.

https://www.cdc.gov/nchs/nhis/index.html

---

**Table 1:**
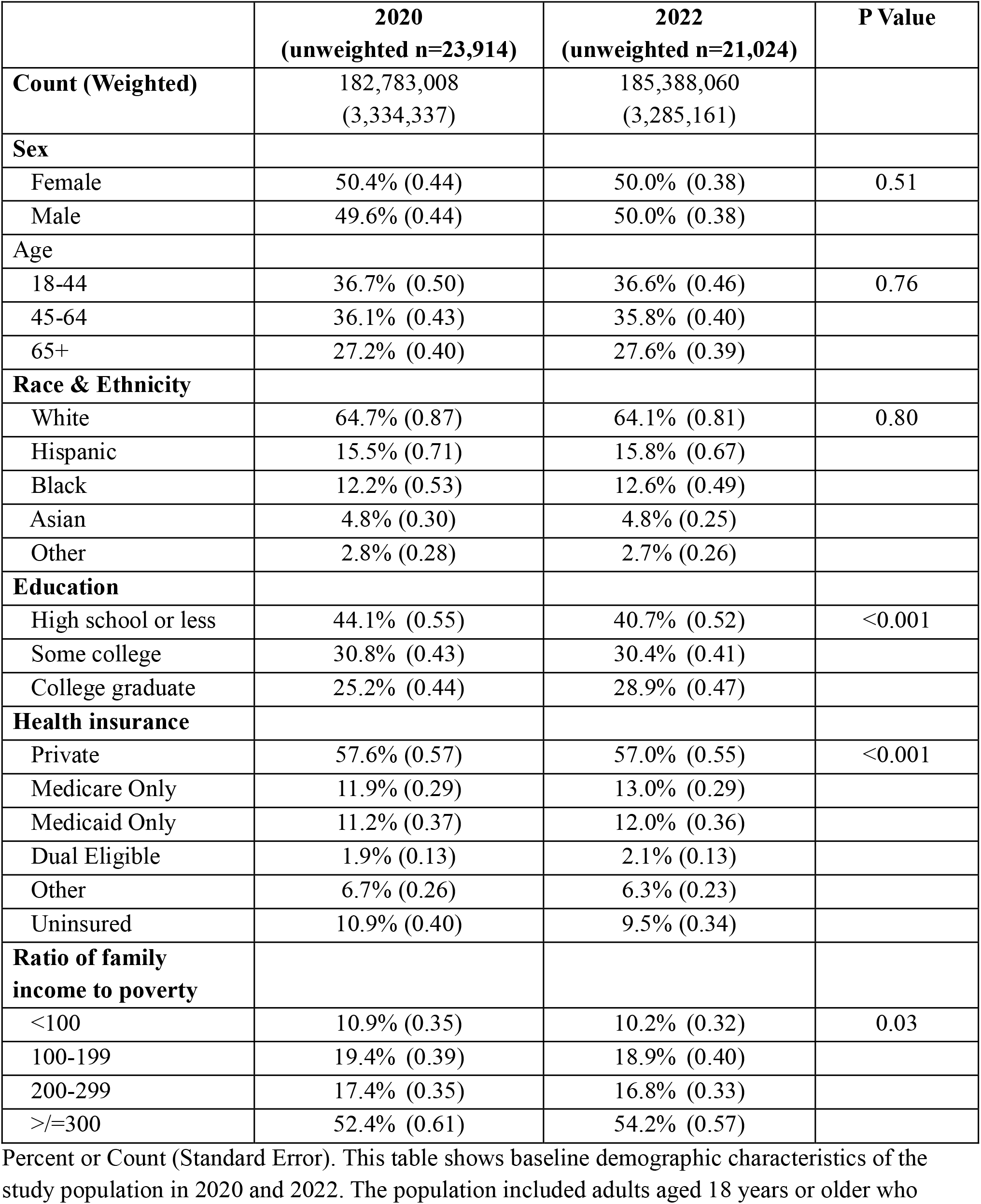
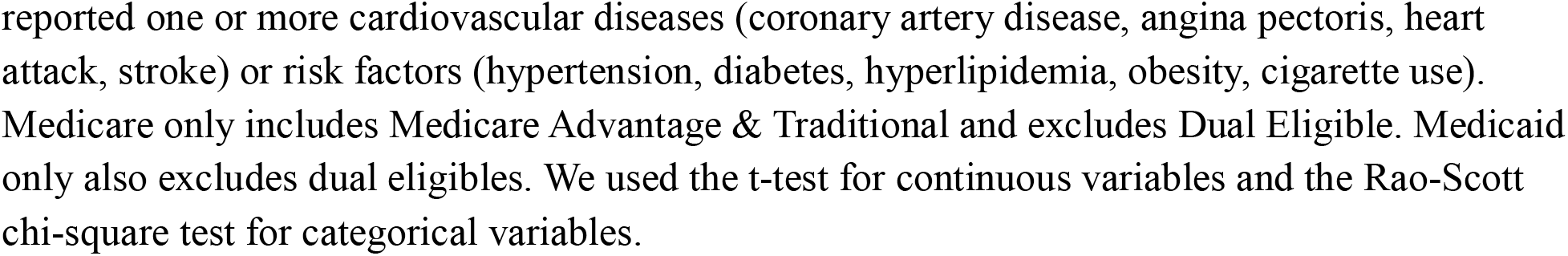
Baseline Characteristics Among US Adults with Cardiovascular Risk Factors or Disease in 2020 and 2022.

**Table: Baseline Characteristics Among US Adults with Cardiovascular Risk Factors or Disease in 2020 and 2022**
|  | **2020**<br>**(unweighted n=23,914)** | **2022**<br>**(unweighted n=21,024)** | **P Value** |
| --- | --- | --- | --- |
| **Count (Weighted)** | 182,783,008<br>(3,334,337) | 185,388,060<br>(3,285,161) |  |
| **Sex** |  |  |  |
| Female | 50.4% (0.44) | 50.0% (0.38) | 0.51 |
| Male | 49.6% (0.44) | 50.0% (0.38) |  |
| **Age** |  |  |  |
| 18-44 | 36.7% (0.50) | 36.6% (0.46) | 0.76 |
| 45-64 | 36.1% (0.43) | 35.8% (0.40) |  |
| 65+ | 27.2% (0.40) | 27.6% (0.39) |  |
| **Race & Ethnicity** |  |  |  |
| White | 64.7% (0.87) | 64.1% (0.81) | 0.80 |
| Hispanic | 15.5% (0.71) | 15.8% (0.67) |  |
| Black | 12.2% (0.53) | 12.6% (0.49) |  |
| Asian | 4.8% (0.30) | 4.8% (0.25) |  |
| Other | 2.8% (0.28) | 2.7% (0.26) |  |
| **Education** |  |  |  |
| High school or less | 44.1% (0.55) | 40.7% (0.52) | <0.001 |
| Some college | 30.8% (0.43) | 30.4% (0.41) |  |
| College graduate | 25.2% (0.44) | 28.9% (0.47) |  |
| **Health insurance** |  |  |  |
| Private | 57.6% (0.57) | 57.0% (0.55) | <0.001 |
| Medicare Only | 11.9% (0.29) | 13.0% (0.29) |  |
| Medicaid Only | 11.2% (0.37) | 12.0% (0.36) |  |
| Dual Eligible | 1.9% (0.13) | 2.1% (0.13) |  |
| Other | 6.7% (0.26) | 6.3% (0.23) |  |
| Uninsured | 10.9% (0.40) | 9.5% (0.34) |  |
| **Ratio of family income to poverty** |  |  |  |
| <100 | 10.9% (0.35) | 10.2% (0.32) | 0.03 |
| 100-199 | 19.4% (0.39) | 18.9% (0.40) |  |
| 200-299 | 17.4% (0.35) | 16.8% (0.33) |  |
| >=300 | 52.4% (0.61) | 54.2% (0.57) |  |

Percent or Count (Standard Error). This table shows baseline demographic characteristics of the study population in 2020 and 2022. The population included adults aged 18 years or older who
reported one or more cardiovascular diseases (coronary artery disease, angina pectoris, heart attack, stroke) or risk factors (hypertension, diabetes, hyperlipidemia, obesity, cigarette use). Medicare only includes Medicare Advantage & Traditional and excludes Dual Eligible. Medicaid only also excludes dual eligibles. We used the t-test for continuous variables and the Rao-Scott chi-square test for categorical variables.

**Figure 1:**
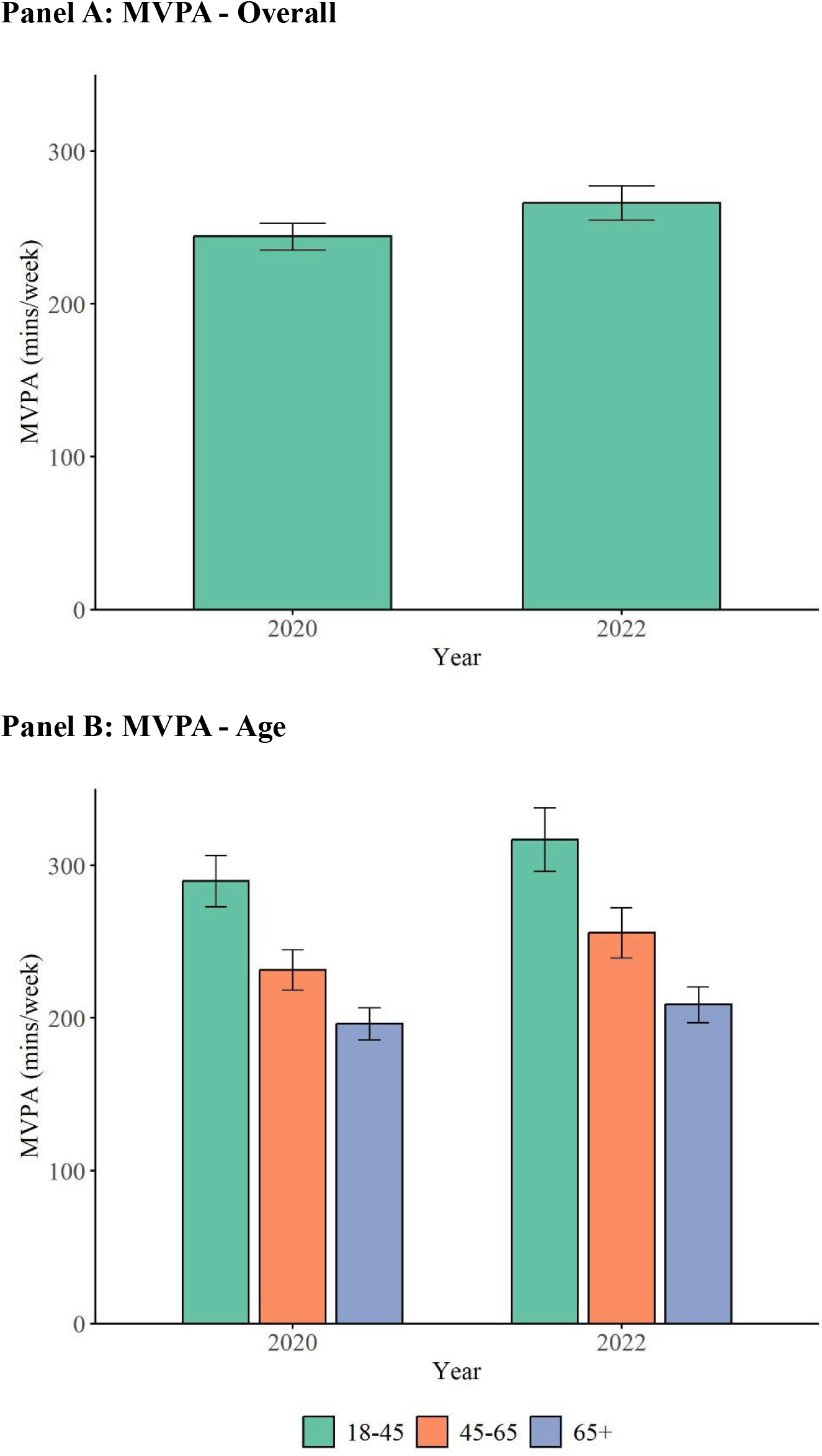
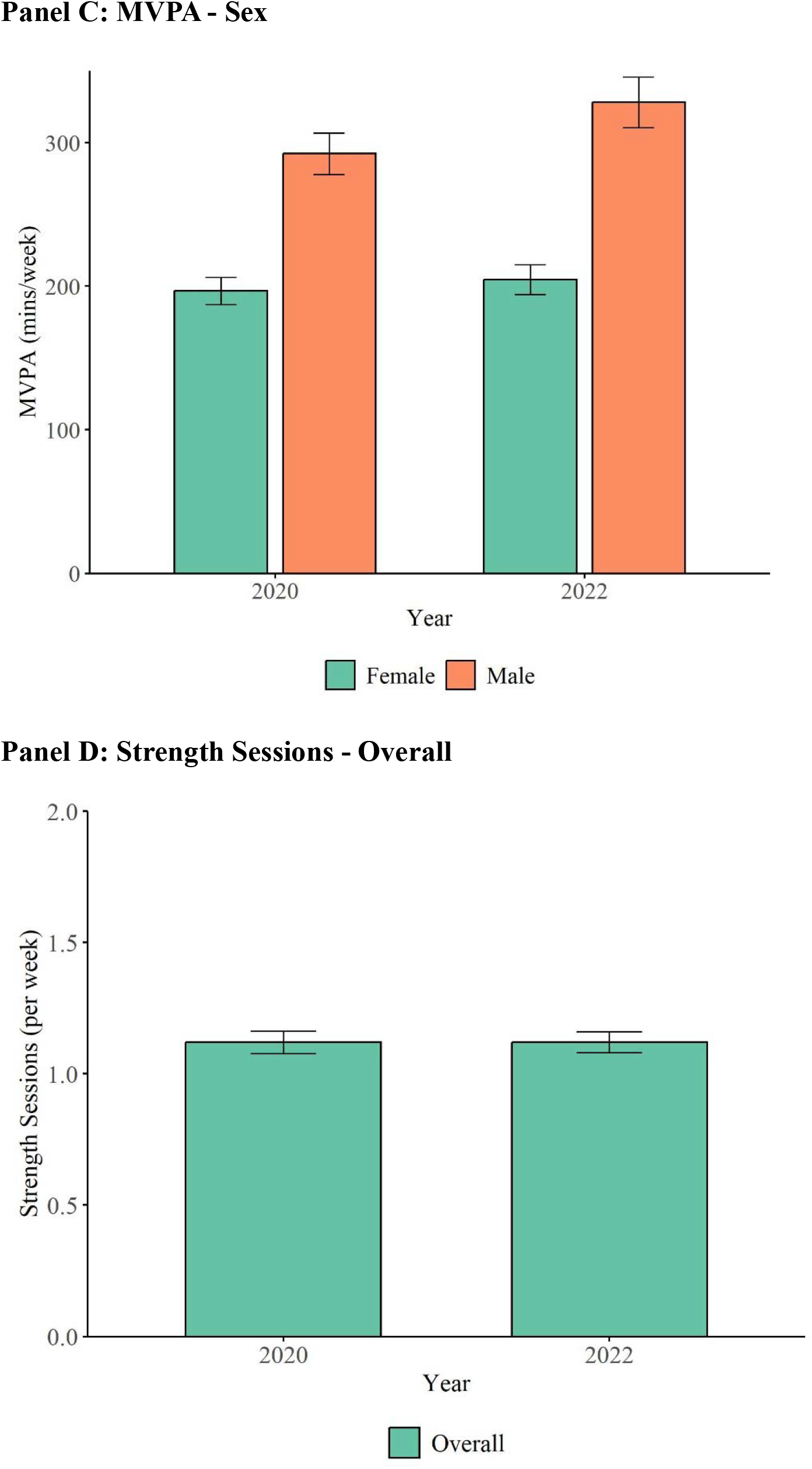
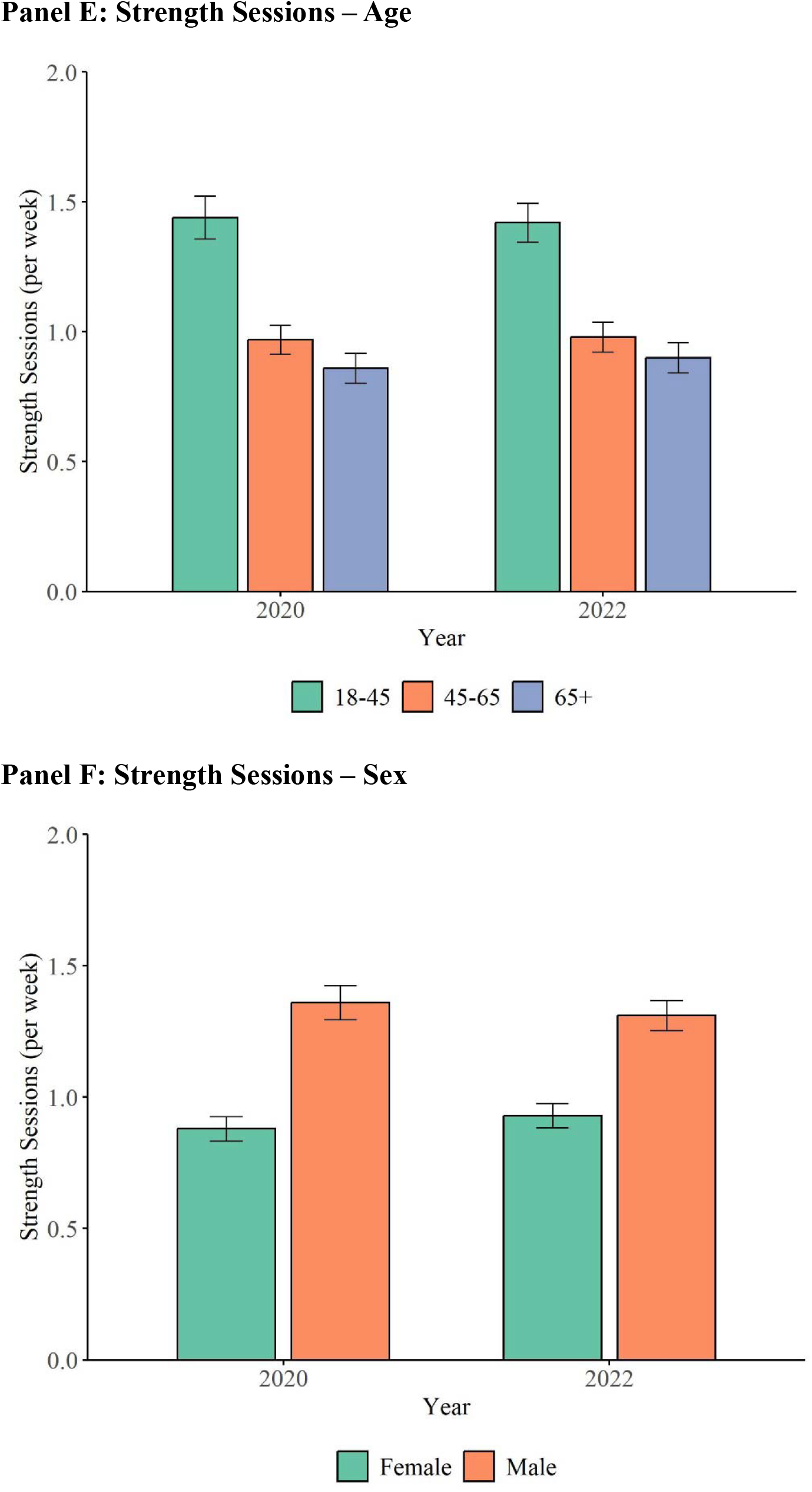
Moderate-to-Vigorous Physical Activity (MVPA) and Strength Sessions Among US Adults with Cardiovascular Risk Factors or Disease, 2020 and 2022, Overall and by Sex and Age.

This figure summarizes mean moderate-to-vigorous physical activity (MVPA) and strength sessions among US adults with cardiovascular risk factors or disease in 2020 and 2022. Panel A represents the results for MVPA for the overall population. Panel B is a subgroup analysis of MVPA by age. Panel C is a subgroup analysis of MVPA by sex. Panel D represents the results for strength sessions for the overall population. Panel E is a subgroup analysis of strength sessions by age. Panel F is a subgroup analysis of strength sessions by sex. Error bars represent 95% confidence intervals for each group mean. Statistical comparisons between 2020 and 2022 are based on adjusted regression models and should not be inferred from visual overlap of CIs.

## References

1. Marinacci LX, Bartlett V, Zheng Z, Mein S, Wadhera RK. Health Care Access and Cardiovascular Risk Factor Management Among Working-Age US Adults During the Pandemic. Circ Cardiovasc Qual Outcomes. Dec 2023;16(12):e010516. doi:10.1161/circoutcomes.123.010516

2. Alba C, Zheng Z, Wadhera RK. Changes in Health Care Access and Preventive Health Screenings by Race and Ethnicity. JAMA Health Forum. 2024;5(2):e235058–e235058. doi:10.1001/jamahealthforum.2023.5058

3. American Time Use Survey News Release: American Time Use Survey --May to December 2019 and 2020 Results. Bureau of Labor Statistics. https://www.bls.gov/news.release/archives/atus_07222021.htm

4. Ng TKY, Kwok CKC, Ngan GYK, et al. Differential Effects of the COVID-19 Pandemic on Physical Activity Involvements and Exercise Habits in People With and Without Chronic Diseases: A Systematic Review and Meta-analysis. Arch Phys Med Rehabil. Jul 2022;103(7):1448-1465.e6. doi:10.1016/j.apmr.2022.03.011

5. Ji H, Gulati M, Huang TY, et al. Sex Differences in Association of Physical Activity With All-Cause and Cardiovascular Mortality. JACC. 2024;83(8):783–793. doi:10.1016/j.jacc.2023.12.019

6. Paluch AE, Boyer WR, Franklin BA, et al. Resistance Exercise Training in Individuals With and Without Cardiovascular Disease: 2023 Update: A Scientific Statement From the American Heart Association. Circulation. 2024/01/16 2024;149(3):e217–e231. doi:10.1161/CIR.0000000000001189

